# Pudendal somatosensory evoked potentials – A standardized assessment for males and females

**DOI:** 10.1101/2025.11.21.25340767

**Authors:** Jason M Anasori, Winston Brandt, Stephanie Wilkinson, Alison MM Williams, Lukas D Linde, John LK Kramer, Tania Lam

## Abstract

**Objective:** To evaluate configuration- and sex-related differences in pudendal somatosensory evoked potential (SEP) waveform characteristics, tolerability, response stability, and test–retest reliability in healthy adults.

**Methods:** Pudendal SEPs were recorded from forty-two able-bodied adults (20 females, 22 males) across multiple electrode configurations that varied in anode-cathode placement for both sexes, and in laterality (unilateral and bilateral) for females only. Tibial SEPs were also recorded as a reference control. Peak-to-peak amplitudes were compared across configurations and between sexes and nerves using linear mixed-effects models. SEP latency was summarized descriptively. Tolerability (self-reported 1–10 Likert scale) was analyzed with cumulative link mixed models. Peak-to-peak amplitude stability was computed from exponential fits of cumulative averages and compared across configurations and nerves. Test–retest reliability for SEP latency and peak-to-peak amplitudes was assessed using intraclass correlation coefficients and Bland–Altman analyses.

**Results:** Electrode configuration had no effect on peak-to-peak amplitude in females or males, and no sex-based amplitude differences were observed. Tolerability varied significantly by configuration, where the configuration with the clitoral cathode and bilateral anodes was rated least tolerable in females, and the configuration with the anode on the glans and the cathode on the proximal shaft was least tolerable in males. Tibial nerve stimulation was consistently rated as more tolerable than pudendal, with a larger difference observed in females. On average, pudendal SEP amplitudes stabilized at 282 stimuli in females and 324 in males, with no differences across configurations nor compared to tibial SEPs. Test–retest analyses showed no systematic bias and revealed moderate-to-good reliability, with latency measures demonstrating greater reproducibility than peak-to-peak amplitudes.

**Conclusions:** Pudendal SEP amplitudes and response stability were robust to electrode configuration, but tolerability differed across configurations, highlighting the need to consider electrode placement when designing protocols to improve participant comfort. Test–retest analyses demonstrated moderate-to-good reproducibility, particularly for latency measures, supporting the reliability of pudendal SEPs for longitudinal or repeated assessments.

**Significance:** These findings establish the feasibility and reliability of pudendal SEPs in healthy adults and provide guidance for optimizing stimulation locations in future research.

## 1.0 Introduction

Bladder, bowel, and sexual function are essential to quality of life, and disruptions in any of these systems can have profound physical, emotional, and social consequences (Aoun et al., 2021; Hibner et al., 2010; Khoder & Hale, 2014; Schraffordt et al., 2004). The pudendal nerve provides the motor, autonomic, and sensory innervation to key pelvic structures associated with each of these functions. It originates from the S2-S4 spinal segments and trifurcates into the inferior rectal nerve, the perineal nerve, and the dorsal nerve of the clitoris or penis (Aoun et al., 2021; Cavalcanti et al., 2007; Fadel et al., 2021; Hibner et al., 2010; Khoder & Hale, 2014; Schraffordt et al., 2004; Shafik et al., 1995). Direct injury to pudendal nerve branches or lesions to the associated ascending and descending spinal pathways may result in a range of functional impairments. Given its importance, accurate assessments of pudendal nerve function are essential for identifying and addressing dysfunction.

Somatosensory evoked potentials (SEPs) provide a means to evaluate pudendal nerve sensory function and ascending tract integrity in various clinical populations. In this assessment, electrical stimuli are delivered to a branch of the pudendal nerve and subsequent cortical responses are captured using electroencephalography (EEG) over the somatosensory cortex (Cruccu et al., 2008; Lüders et al., 1985; Muzyka & Estephan, 2019). In previous research, the majority of protocols have targeted the dorsal nerve of the penis or clitoris to elicit pudendal SEPs, with considerable variability in the configuration and location of stimulating electrodes (Williams et al., 2024). In females, surface electrodes are commonly affixed along the vulva near the clitoris and labia in either a unilateral or bilateral configuration; in males, the most common stimulation sites are along the dorsal penile shaft or glans penis (Williams et al., 2024). Stimulus intensity is typically set to 2-4 times the perceptual threshold (Williams et al. 2024).

One of the factors that contributes to participant burden is the number of stimuli delivered. Pudendal nerve stimulation can be uncomfortable, and delivering hundreds of stimuli may amplify this discomfort. Clinical recommendations for SEPs suggest averaging at least 500 trials to ensure reproducibility and accurate measurement of latency and amplitude (Cruccu et al., 2008). However, in a recent systematic review, Williams et al. (2024) reported that the median number of stimuli used in previous studies recordings pudendal SEPs was 300, with a range from 50 to 2048, suggesting considerable variability across studies. To balance participant tolerability with the need for high-quality data, it is important to determine the minimum number of stimuli required to produce a stable SEP waveform.

Waveform characteristics have been used to evaluate pudendal nerve function before and after an intervention or procedure (Calabrò et al., 2019; Dasgupta et al., 2004; Giani et al., 2011; Ma et al., 2019; Senol et al., 2008; Song et al., 2020; Xia et al., 2016). The most commonly reported pudendal SEP characteristics are latency and peak-to-peak amplitude of specific waveform features (Williams et al., 2024). However, the between-day reliability of these measures in healthy adults and clinical populations has not yet been established for pudendal SEPs. In contrast, studies assessing SEPs from other peripheral nerves have shown variable reproducibility across nerves and recording conditions. Ehrenbrusthoff et al. (2022) developed a SEP protocol targeting sensory nerves of the lower back and reported very poor test–retest reliability in healthy adults, with large random variability between sessions. Hardmeier et al. (2019) noted that relatively few SEP reliability investigations exist but reported consistent cortical latencies for median-nerve SEPs in a multicentre sample of individuals with multiple sclerosis. Similarly, Brown et al. (2017) found that SEPs elicited from the median nerve were among the most reliable electrophysiological measures across study sites. Establishing the test-retest reliability of pudendal SEP outcomes is therefore critical, particularly if these measures are to be used for longitudinal tracking of patients or evaluating the effects of therapeutic interventions.

Despite the potential importance of pudendal SEPs in clinical and research settings, techniques used to elicit and record pudendal SEPs remain inconsistent in the literature, best practice guidelines have yet to be established, and the reliability of testing outcomes has not yet been examined. Furthermore, eliciting pudendal SEPs is further complicated by the multiple possible configurations of electrode placement within and between females and males, unlike other peripheral nerves commonly used for SEP assessment (e.g. median nerve, tibial nerve) where the anatomical location for delivering electrical stimulation is more straightforward. Thus, the primary aim of this study was to determine if there are configuration-dependent effects on pudendal SEP waveform characteristics, tolerability to stimulation, and the number of stimuli required to achieve a stable pudendal SEP response in female and male adults. Secondary aims of this study were to 1) determine if there were any sex-based differences in pudendal SEP waveform characteristics, tolerability to pudendal stimulation, and pudendal SEP response stability; and 2) evaluate the test-retest reliability of pudendal SEP waveform characteristics.

## 2.0 Methods

### 2.1 Participants

We recruited able-bodied adults aged 19 years and older for this study. We excluded participants if they had a neurological impairment; reported symptoms of urinary, bowel, or sexual dysfunction; were pregnant or had been pregnant within the past 6 months; had previous surgery involving the sex organs or perineum (except for circumcision in males performed at least 6 months earlier); or had a genital piercing. Ethical approval was obtained from the University of British Columbia’s Clinical Research Ethics Board, and all participants provided written informed consent.

### 2.2 Electrode Configurations

In female and male participants, we stimulated the dorsal nerve of the clitoris or penis, respectively. Figure 1 outlines the electrode configurations for each sex.

**Figure 1.**
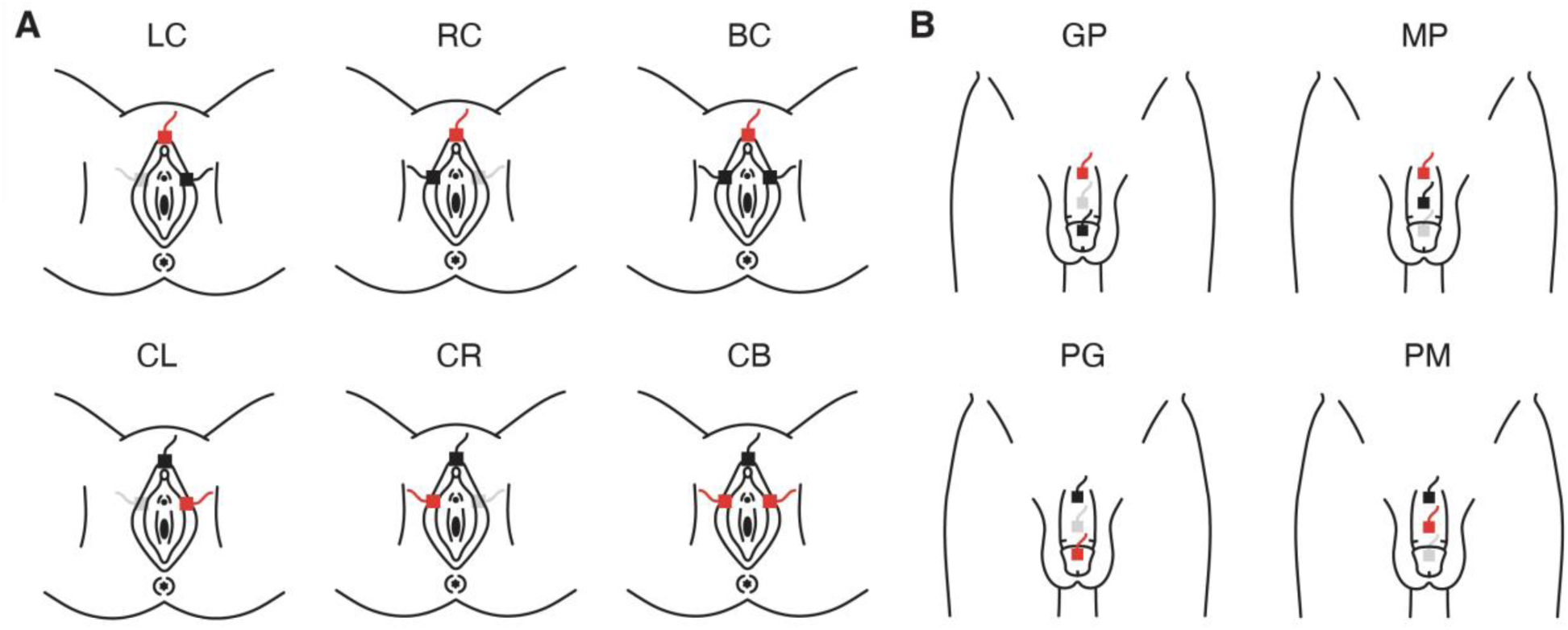
Electrode configurations for pudendal nerve stimulation in females (A) and males (B). In females, we tested six configurations, and in males, we tested four configurations. Red electrodes indicate cathodes, and black electrodes indicate anodes. Grey electrodes represent inactive sites in a given configuration. Configuration abbreviations denote electrode polarity and location, with the first letter representing the anode position and the second the cathode. In males, G = glans, P = proximal shaft, M = mid-shaft. In females, R = right, L = left, B = bilateral, and C = clitoris.

For female participants, three 12-mm square disposable Ag/AgCl surface electrodes were applied: one on the clitoral hood and two positioned within the crease between the outer and inner labia at the level of the urethra. We tested six female stimulation configurations. Four configurations used two electrodes to stimulate unilaterally, and two configurations involved all three electrodes to stimulate bilaterally (Fig. 1A).

For male participants, three 2.5-cm disposable cloth surface electrodes were placed on the dorsal side of the penis: one electrode on the glans, and two electrodes positioned 2-3 cm apart along the midline of the shaft. We tested four male stimulation configurations, which always paired the proximal shaft electrode with either the distal shaft or glans electrode, while alternating the location of the anode (Fig. 1B).

We also recorded tibial SEPs as a control condition in female and male participants. The tibial nerve was stimulated via two 2.5-cm disposable cloth surface electrodes affixed near the ankle, posterior to the medial malleolus, with the cathode placed proximally and the anode placed distally.

### 2.3 Recording Procedures

Participants were positioned comfortably in supine on a plinth in a dimly lit room. We recorded EEG signals from Cz and referenced to Fz using a 32-channel EEG system with a sampling frequency of 5 kHz (actiCHamp Plus, Brain Vision Solutions, Montreal, QC, Canada).

For each pudendal and tibial nerve electrode configuration, we identified the participant’s perceptual threshold (PT). This was done by slowly increasing the stimulus intensity until the participant reported that they could first identify the rhythm of the continuous 3 Hz stimulus train. We then raised the intensity to approximately 120% of this value and gradually reduced the intensity until the participant reported no longer detecting the sensation. We repeated this ascending-descending procedure three times, recording the intensity at which perception began (up) and ended (down) during each trial. This gave us six values per configuration, and we calculated PT as the mean of those six values. Table 1 summarizes the mean and range of the current amplitude at PT for each configuration.

**Table 1.** Perceptual threshold (milliamp, mA), reported as mean ± standard deviation (range).

| Configuration | PT (mA) |
| --- | --- |
| **Female** |  |
| CB | 2.1 ± 0.9 (0.5 - 3.8) |
| BC | 2.3 ± 1.0 (1.1 - 4.2) |
| CL | 1.7 ± 0.6 (0.3 - 2.7) |
| LC | 2.0 ± 0.8 (0.7 - 3.7) |
| CR | 1.7 ± 0.7 (0.5 - 2.8) |
| RC | 2.1 ± 0.9 (0.7 - 3.8) |
| Tibial | 1.1 ± 0.5 (0.6 - 2.9) |
| **Male** |  |
| PG | 2.5 ± 0.9 (1.2 - 4.5) |
| GP | 2.8 ± 1.2 (1.4 - 5.2) |
| PM | 2.3 ± 1.0 (1.2 - 5.6) |
| MP | 2.9 ± 1.4 (1.2 - 5.5) |
| Tibial | 2.0 ± 0.7 (1.0 - 3.4) |

We recorded pudendal and tibial SEPs using a stimulus intensity of 3×PT. If a participant could not tolerate 3×PT, we lowered the stimulus intensity to the highest tolerable intensity. If a participant could not tolerate at least 2×PT for a given configuration, we excluded the configuration for this participant. We excluded 17 trials across 5 participants due to intolerance of stimulus intensity above 2×PT (Fig. 3).

For each pudendal and tibial nerve stimulation configuration, we delivered 600 square wave pulses of 1 millisecond duration using a constant-current stimulator (DS7R, Digitimer, Welwyn Garden City, Hertfordshire, UK) at randomized frequencies ranging from 1 to 3 Hz. We randomized the order of electrode configuration tested for each participant, with the exception of the tibial nerve stimulation trial, which was always the middle trial of the session. After each trial, participants gave a tolerability score for the given configuration on a scale of 1 to 10, where 1 represented barely perceivable stimulation and 10 represented unbearable intensity.

### 2.4 Data Analysis

We analyzed our data using the EEGLAB toolbox (Delorme & Makeig, 2004) for MATLAB R2024b (MathWorks, Natick, MA, USA) and custom MATLAB scripts. All EEG data were preprocessed using high-pass and low-pass filters with cut-off frequencies of 1 Hz and 100 Hz, respectively. We averaged the time-locked EEG signals of the 600 stimuli, and two of the authors (SW, WB) independently reviewed the traces to identify the presence and location of each peak (P1, N1, P2, N2). Discrepancies greater than 0.5 ms in peak latency were resolved through consensus with a third author (AMMW). For waveforms with identifiable peaks, we extracted the latency and peak-to-peak amplitude of each waveform complex (P1N1, N1P2, P2N2) by taking the absolute difference between peaks. Waveforms with no identifiable peaks did not contribute to further analysis.

To determine when the SEP stabilized, the peak-to-peak amplitude derived from the 600-stimuli average was compared to amplitudes of incremental averages, starting with a 2-stimuli average. The standard deviation of the peak-to-peak amplitude for each complex was calculated, and an exponential line of best fit was applied using Equation 1 to determine the time constant, τ, representing 63.2% of the final stable value. The stabilization point (τ_95_) was then calculated using Equation 2, corresponding to the number of stimuli at which the standard deviation settled to 95% of its final value. The peak-to-peak amplitude for each complex was subsequently calculated using averaged responses up to τ_95_ for each trial, or the τ_95_-derived peak-to-peak (P2P_τ95_) and compared to the average peak-to-peak amplitude from the whole trial containing 600 stimuli (P2P_600_). Only trials where the exponential line of best fit was good (R^2^ ≥ 0.80) and τ_95_ ≤ 2048 stimuli were included (2048 was the highest number of stimuli delivered across different pudendal SEP protocols reported in the literature) (Williams et al., 2024).

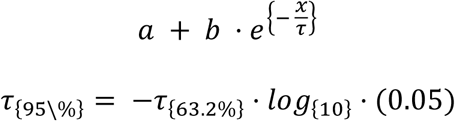

Figure 2 presents a representative example of response stability analysis for pudendal SEPs, showing the cumulative P2P_600_ curve and the standard deviation–based fit used to determine τ₉₅.

**Figure 2.**
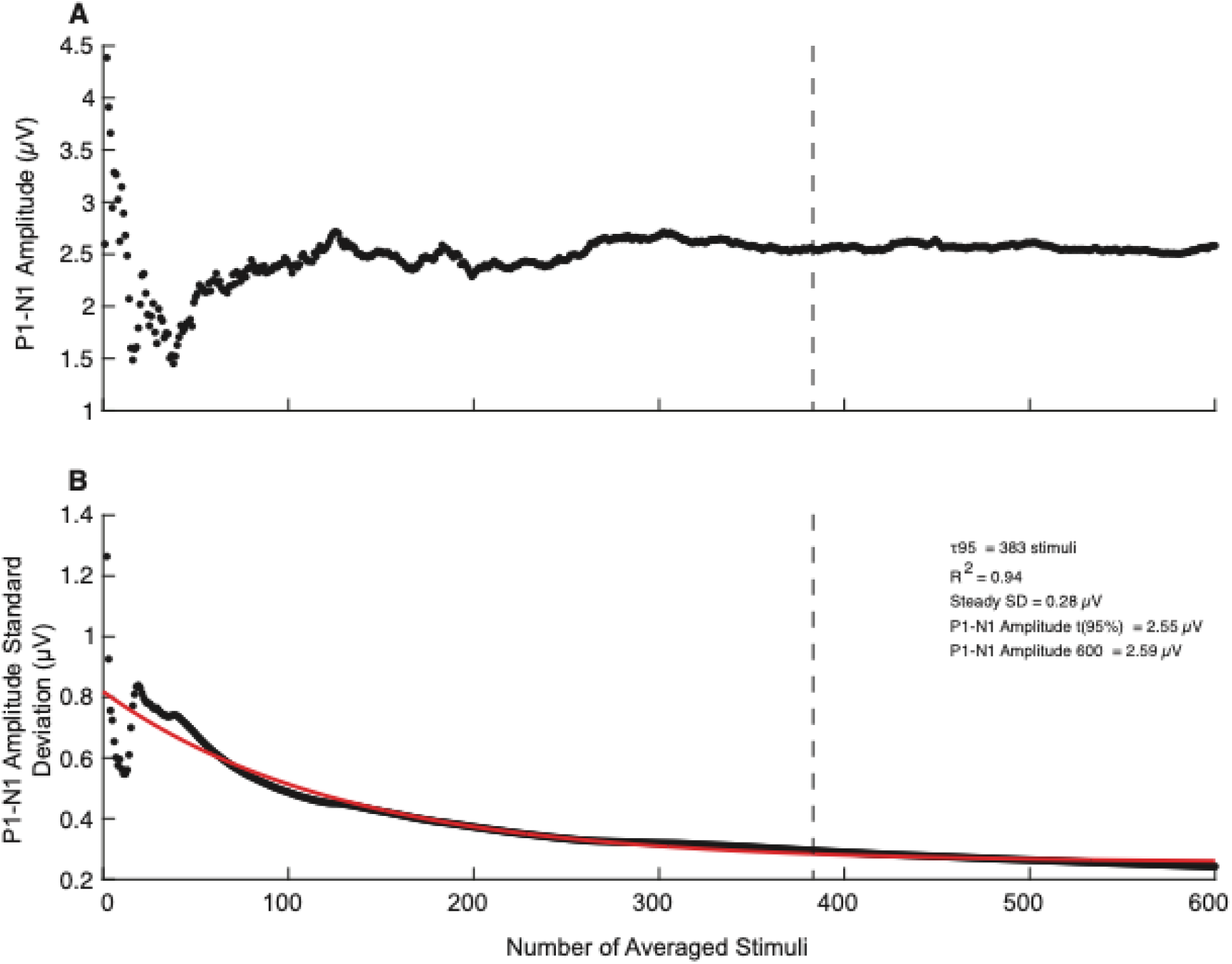
Representative example of response stability analysis for pudendal SEPs (P1N1 complex). The cumulative P2P₆₀₀ curve is plotted against the number of averaged stimuli, with the vertical dashed line marking the τ₉₅ stabilization point (A). The standard deviation of P1N1 amplitude across incremental averages with exponential fit is shown below, from which P2Pτ₉₅ is calculated (B).

### 2.5 Statistical Analysis

All analyses were conducted using RStudio (version 2024.12.1+563; R Foundation for Statistical Computing, Vienna, Austria) and statistical significance was set to 0.05. The number of identifiable peaks (P1, N1, P2, N2) and the latencies of the peaks were summarized using descriptive statistics. To compare the effect of electrode configuration on peak-to-peak amplitudes (P1N1, N1P2, P2N2), we used linear mixed-effects models (LMERs) using the lmer function from the lme4 package (Bates et al., 2015), with p-values obtained via the lmerTest package (Kuznetsova et al., 2017) using Satterthwaite’s method. Peak-to-peak amplitudes of each complex (P1N1, N1P2, P2N2) were modelled as dependent variables. Electrode configuration was included as a fixed effect, and participants were included as a random intercept to account for repeated measures. Since female and male pudendal electrode configurations were not comparable, we performed separate LMERs for each group. The reference electrode configuration was CB for females (clitoral anode, bilateral cathodes) and GP for males (glans anode, proximal shaft cathode). GP was chosen for males as it reflects the most commonly used configuration in previous pudendal SEP literature (Williams et al., 2024). CB was selected for females because it engages both left and right pudendal branches simultaneously, with the anode on the clitoris (analogous to the male GP configuration). We log-transformed the peak-to-peak amplitude data prior to analysis to meet assumptions of normality and homogeneity of variance. Model assumptions for LMERs were assessed using residual and Q-Q plots, along with formal diagnostic tests from the performance (Lüdecke et al., 2021) and DHARMa packages (Hartig, 2022). These included checks for normality (via simulated residuals and uniformity tests) and overdispersion. If the model revealed a significant main effect, we conducted post-hoc pairwise comparisons between configurations using estimated marginal means. To account for multiple comparisons, we applied the Bonferroni correction to adjust the p-value based on the total number of possible pairwise contrasts conducted across three separate models. For females, there are 15 possible pairwise comparisons across 6 configurations, so we adjusted p-values based on a total of 45 possible comparisons. For the male data, there are 6 possible pairwise comparisons across 4 configurations, so we adjusted p-values based on a total of 18 possible comparisons.

To determine whether peak-to-peak amplitudes differed between sexes, we first averaged the amplitude across all electrode configurations for each participant. We then performed independent t-tests comparing females and males for each SEP complex. To correct for multiple comparisons across the three complexes, we used a Bonferroni-adjusted α of 0.017 (0.05/3).

To evaluate differences in tolerability scores across pudendal configurations, we used cumulative link mixed models (CLMMs), which are appropriate for ordinal data. We fit the female and male data separately using the clmm function from the ordinal package (Christensen, 2019), with tolerability scores modelled as a function of pudendal electrode configuration and participants included as a random intercept. Tibial trials were excluded from these models. For females, we set the reference to BC instead of CB, due to reduced variability and clustering in CB tolerability scores. For males, we set the reference configuration to GP to align with our LMER analysis. If the model revealed a significant main effect, we conducted post hoc pairwise contrasts using estimated marginal means. To account for multiple comparisons, we Bonferroni-adjusted p-values based on the number of possible pairwise contrasts across the different electrode configurations as described above (6 comparisons for males and 15 for females).

To evaluate differences in tolerability scores based on nerve type (pudendal (averaged across all configurations) vs tibial) and sex (female vs male), we fit a combined CLMM. We modelled tolerability scores as a function of nerve type, sex, and their interaction, with a random intercept for participants to account for repeated measures. We selected tibial as the reference level for nerve type, as it was the only configuration common in both sexes, and male was set as the reference level for sex. We directly compared tolerability scores between tibial and pudendal stimulation within each sex using post hoc pairwise tests. To correct for multiple comparisons, we multiplied the resulting p-values by 2 to account for the two primary contrasts of interest.

To determine the validity of our method to establish response stability (τ_95_), we used a paired t-test for each SEP complex to compare the P2P_600_ and P2P_τ95_ amplitudes. Only trials where τ_95_ ≤ 600 stimuli could be included in this part of the analysis. As above, we log-transformed the P2P_τ95_ data to meet assumptions of normality and homogeneity of variance. To correct for multiple comparisons across the three SEP complexes per sex, we used a Bonferroni-adjusted α of 0.017 (0.05/3).

We then evaluated configuration-dependent effects on response stability by modelling τ_95_ following the design of the LMER models described above. For this analysis, we included trials where τ_95_ ≤ 2048. If the model revealed a significant main effect, we conducted post hoc pairwise contrasts using estimated marginal means. To account for multiple comparisons, we applied a Bonferroni correction as described above, to account for 3 complexes x 15 possible pairwise comparisons in females and 3 complexes x 6 comparisons in males.

We also explored the response stability of pudendal SEP responses compared to that of tibial SEP responses by paired t-tests to evaluate differences in τ_95_ across stimulation sites for each complex. Only trials where τ_95_ ≤ 2048 stimuli were included in this analysis. To correct for multiple comparisons across the different complexes, we used a Bonferroni-adjusted α of 0.017 (0.05/3).

To evaluate the reliability of the pudendal SEP procedure, we conducted a test-retest analysis on the latencies and peak-to-peak amplitudes. We averaged waveform observations across configurations to ensure each participant contributed one set of paired data per measure. To determine the reliability of latencies and amplitudes between an initial test and retest, we used an absolute-agreement, single-measure two-way mixed-effect intraclass coefficient (ICC_3,1_) model. We interpreted ICC results via reference to conservative ratings presented by Koo and Li (2016). Bland-Altman confidence values and limits of agreement (mean difference ± 2SD) were calculated, and the mean differences tested against 0 (one-sample t-test) to evaluate bias between sessions (Bland & Altman, 1986).

## 3.0 Results

Forty-three individuals (20 females, 23 males) enrolled in this study. One male participant withdrew from the study due to discomfort with the electrical stimulation. The remaining 42 participants were included in all subsequent analyses. The mean age of the females was 25.3 years (range: 19-36) and males 25.7 years (range: 20-43). All females were nulliparous, and 12 males were circumcised.

### 3.1 Waveform detection

Figure 3 summarizes the detectability of the pudendal SEP peaks (P1, N1, P2, N2) across all electrode configurations in every participant. The detectability of tibial SEP responses is included as a reference. Among the females, there were 13 trials from seven participants with undetectable P1 and N1 peaks, 15 trials from eight participants with undetectable P2 peaks, and 26 trials from 12 participants had undetectable N2 peaks. In the males, there were 10 trials from four participants with undetectable P1 peak, 8 trials from three participants with undetectable N1 and P2 peaks, and 13 trials from eight participants with undetectable N2 peak. Overall, visual inspection of Figure 3 suggests that the variability across participants, and the proportion of trials that had to be excluded either for insufficient stimulus intensity or lack of identifiable waveform, was greater in the female participants (n=65) than the males (n=39).

**Figure 3.**
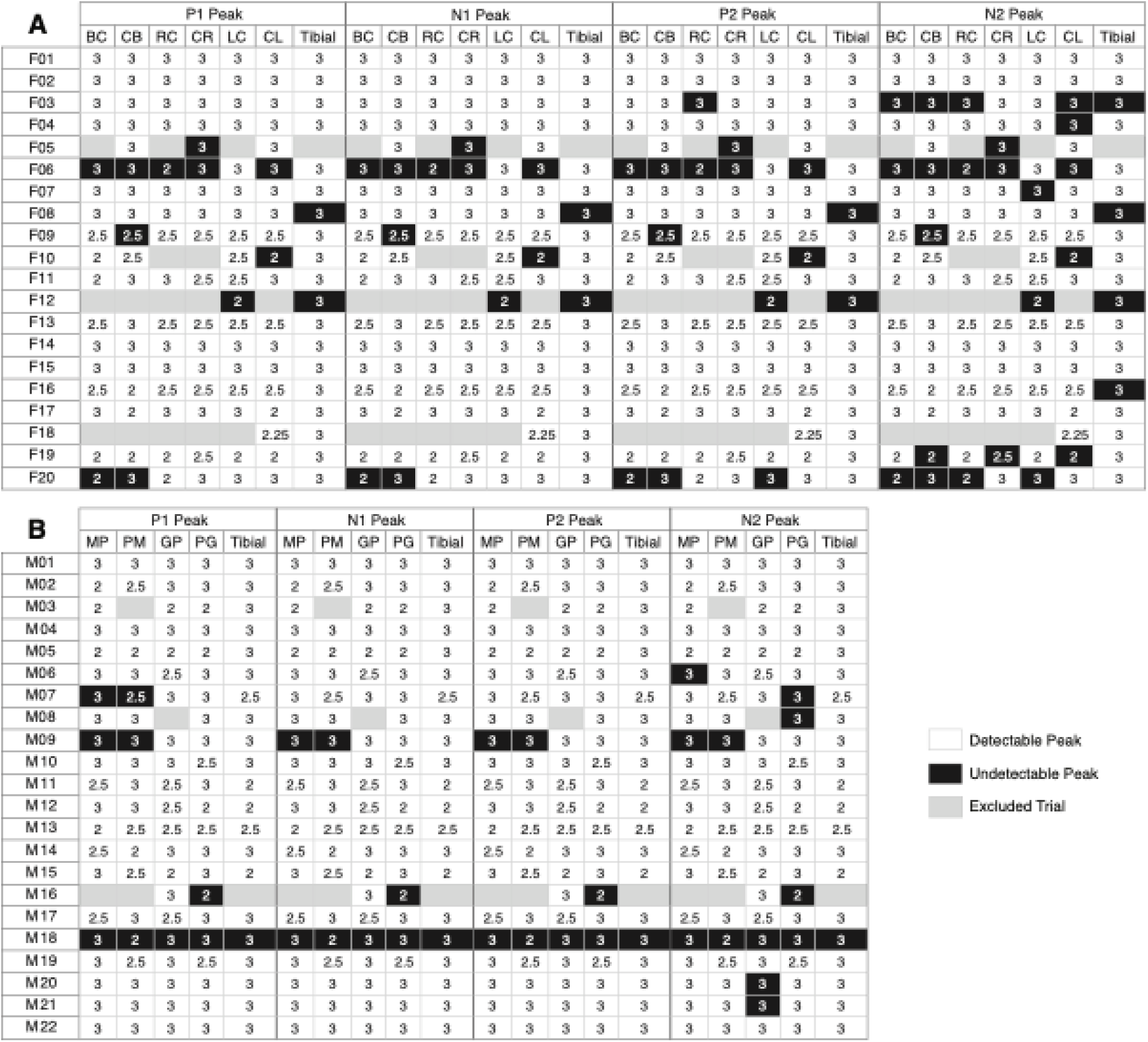
Detectability of SEP peaks (P1, N1, P2, N2) across pudendal and tibial nerve stimulation configurations in female (A) and male (B) participants. Each cell displays the xPT value for a given configuration and peak. White cells indicate detectable peaks; black cells indicate undetectable peaks; grey cells with horizontal lines indicate excluded trials (xPT < 2 or xPT > 3).

### 3.2 Latency

Across all pudendal configurations in all participants, the mean latency of P1 was 41.3 ms (SD: 3.0; range: 32.0-50.0 ms), N1 was 52.4 (SD: 3.4; range: 44.2-62.4 ms), P2 was 64.1 ms (SD: 3.4; range: 55.4-72.4 ms), and N2 was 77.5 ms (SD: 4.0; range: 69.6 - 91.6 ms). In comparison, the mean tibial SEP P1 latency was 44.2 ms (SD: 4.0; range: 36.8–52.0 ms), N1 was 54.5 ms (SD: 4.8; range: 45.0–66.2 ms), P2 was 66.0 ms (SD: 5.2; range: 55.0–76.2 ms), and N2 was 80.0 ms (SD: 5.7; range: 67.0–91.4 ms). The latencies of all the pudendal and tibial SEP waveform components grouped by electrode configuration and sex are presented in Table 2.

**Table 2.**
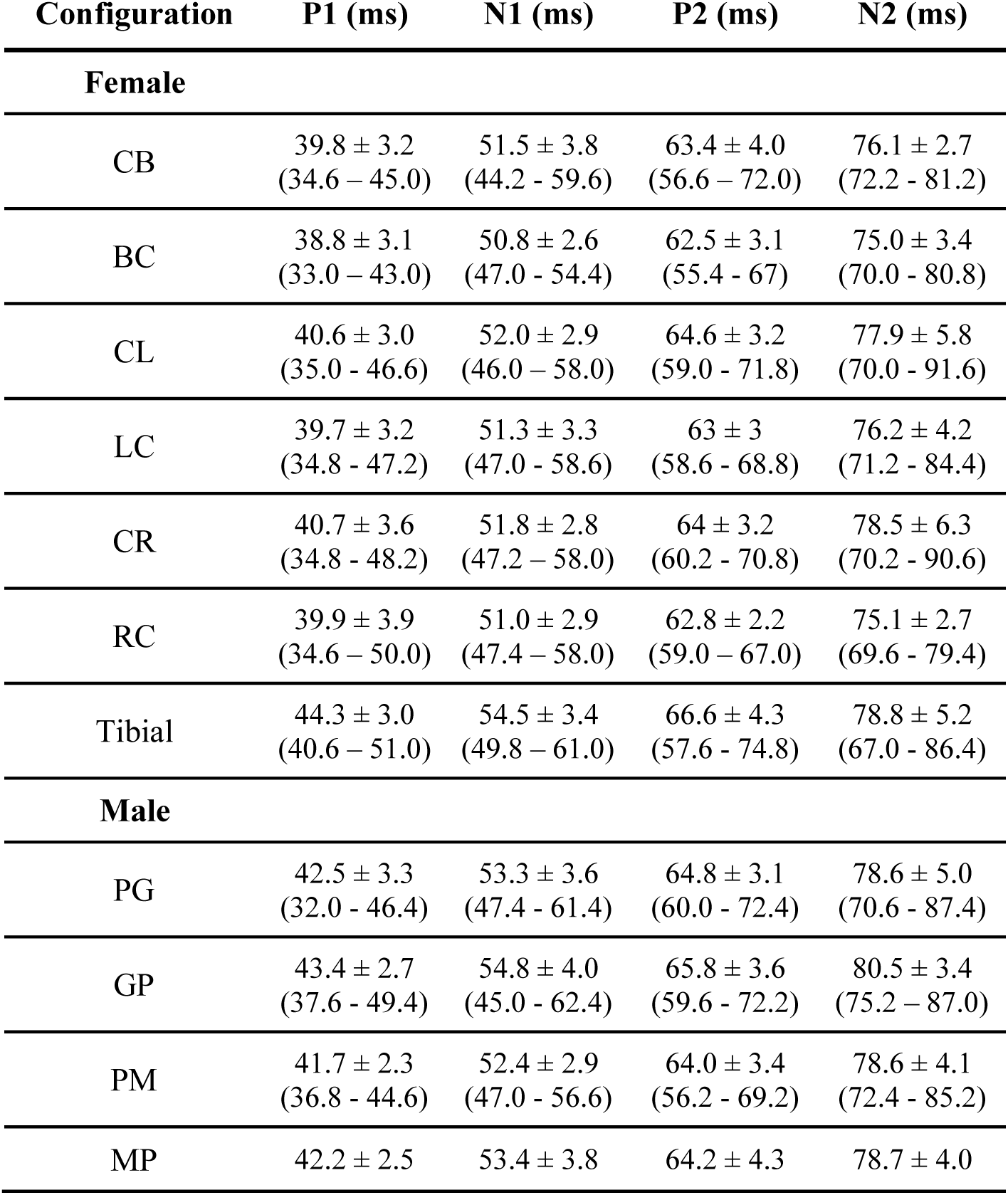
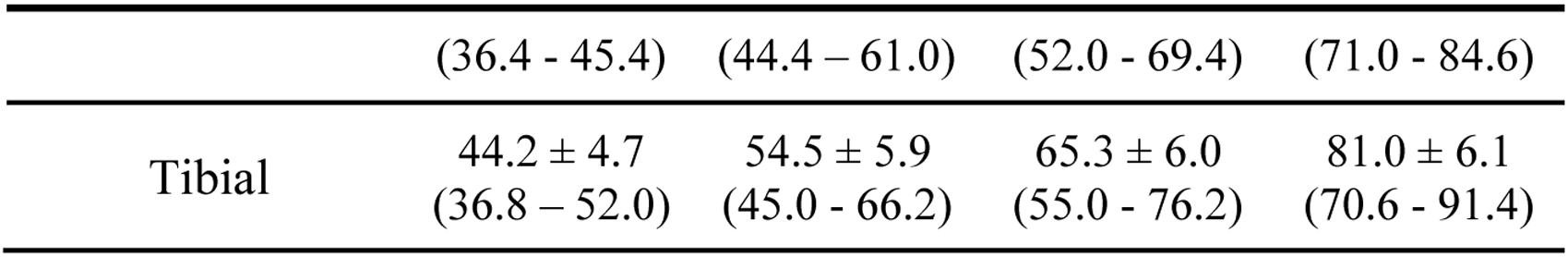
Latency (ms) of each SEP peak, reported as mean ± standard deviation (range).

### 3.3 Effect of electrode configuration on pudendal SEP waveforms

Figure 4 plots the peak-to-peak amplitude of each pudendal SEP waveform complex in females and males. For the P1N1 complex, the LMER analysis revealed no significant effect of electrode configuration on SEP peak-to-peak amplitude in female participants (*p* ≥ 0.2; marginal R² = 0.02; f² = 0.02; Fig. 4A). The mean P1N1 amplitude across all electrode configurations in females was 1.3 μV (SD: 1.0; range: 0.1–5.1 μV). There was also no significant effect of electrode configuration among the male participants (*p* ≥ 0.6; marginal R² = 0.01; f² = 0.01; Fig. 4D). The mean P1N1 amplitude across all electrode configurations in males was 1.1 μV (SD: 1.0; range: 0.1–5.0 μV).

**Figure 4.**
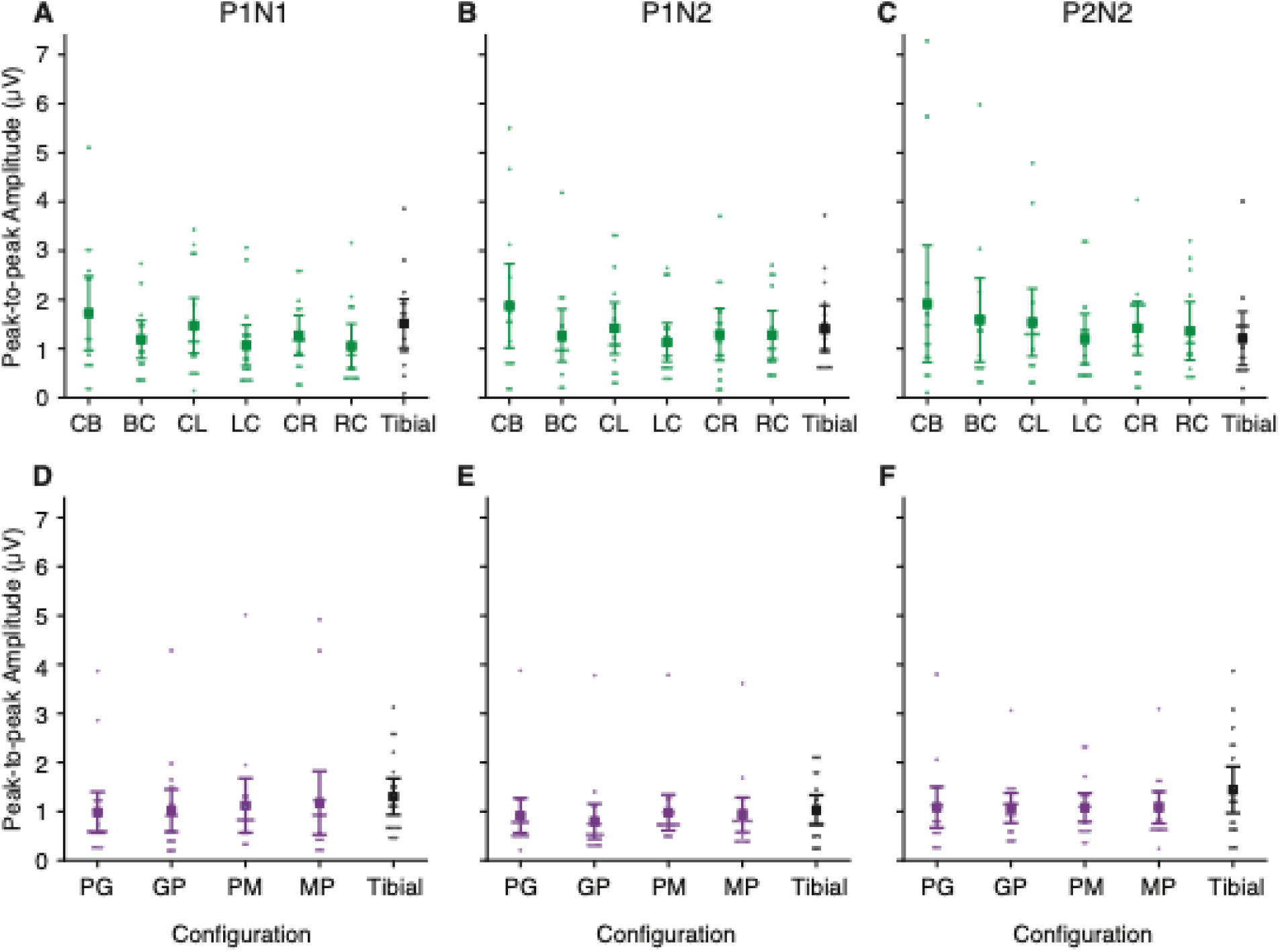
Mean SEP peak-to-peak amplitudes and 95% confidence intervals for each pudendal and tibial configuration in female (top panel) and male (bottom panel) participants. Each column shows the distribution of P1N1, N1P2, and P2N2 complex amplitudes across all stimulation configurations.

There was also no significant effect of electrode configuration on the N1P2 complex in females (*p* > 0.07; marginal R² = 0.02; f² = 0.02; Fig. 4B) or males (*p* > 0.05; marginal R² = 0.03; f² = 0.03; Fig. 4E) and no effect of electrode configuration on the P2N2 complex in either females (*p* ≥ 0.3; marginal R² = 0.01; f² = 0.01; Fig. 4C) or males (*p* > 0.4; marginal R² = 0.003; f² = 0.003; Fig. 4F). The mean amplitude of the N1P2 complex across all electrode configurations was 1.1 μV (SD: 1.0; range: 0.2–5.5 μV) in females and 0.9 μV (SD: 0.7; range: 0.2–3.9 μV) in males. The mean amplitude of the P2N2 complex was 1.5 μV (SD: 2.3; range: 0.1–7.3 μV) in females and 1.2 μV (SD: 0.8; range: 0.2–3.9 μV) in males.

There were no sex-based differences observed for any complex (P1N1 complex: Δ = -0.1, t(32.5) = 0.6, *p* = 1.0, *d* = 0.2); N1P2 complex: Δ = -0.2, t(27) = 1.1, *p* = 0.8, *d* = 0.4); P2N2 complex: Δ = -0.1, t(29.7) = 0.4, *p* = 1.0, *d* = 0.1), all showing comparable peak-to-peak amplitudes between females and males.

### 3.4 Tolerability to protocol

Figure 5 presents the tolerability scores for females and males for each pudendal configuration and tibial nerve stimulation. The mean tolerability score across all pudendal configurations was 4.3 (SD: 1.9; range: 1-9) in females and 4.1 (SD: 1.7; range: 1-8) in males. As a comparison, the mean tolerability score to tibial nerve stimulation was 2.5 (SD: 1.3; range: 1-5) in females and 3.6 (SD: 1.9; range: 1-8) in males.

**Figure 5.**
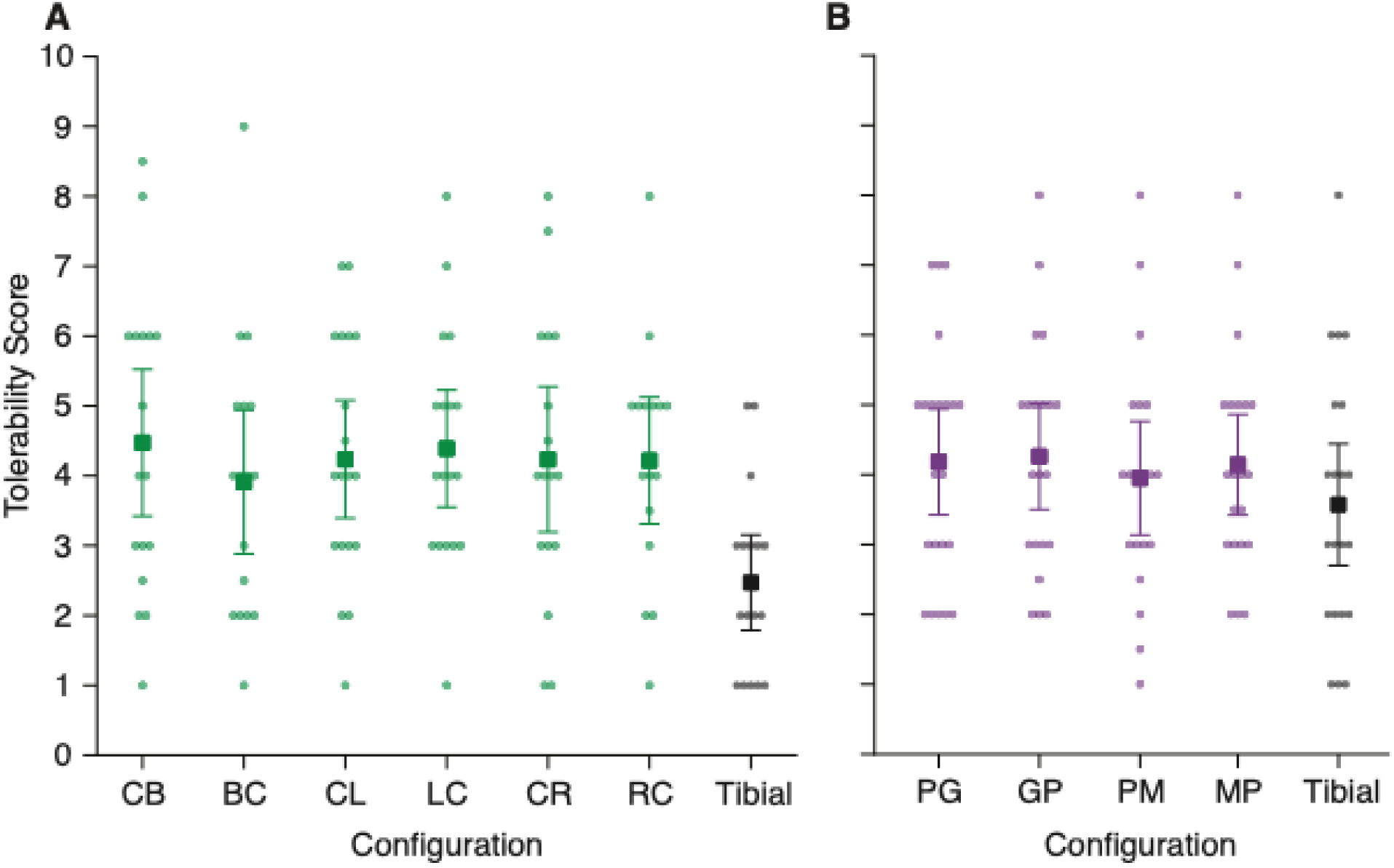
Mean tolerability scores and 95% confidence intervals across pudendal nerve stimulation configurations for females (A) and males (B). Tolerability was rated on a scale from 1 (most tolerable) to 10 (least tolerable).

CLMMs revealed significant differences in tolerability scores across pudendal nerve stimulation configurations for both female and male participants. In females, all pudendal configurations were rated as significantly more tolerable than CB. Post hoc pairwise comparisons indicated that most configuration pairs differed significantly from each other (*p* < 0.001), with the CB configuration being rated least tolerable overall (Table 3).

**Table 3.** Pairwise CLMM contrasts comparing tolerability scores between pudendal configurations in female participants. Each cell shows the odds ratio (row vs. column), 95% confidence interval, and p-value. Odds ratios >1 indicate higher odds of reporting higher tolerability scores (less tolerable) for the row configuration relative to the column configuration.

| Contrast | BC | CL | LC | CR | RC |
| --- | --- | --- | --- | --- | --- |
| **CB** | 4.3<br>[4.25–4.29]<br><0.001 | 2.7<br>[2.67–2.70]<br><0.001 | 1.4<br>[1.38–1.39]<br><0.001 | 4.9<br>[4.84–4.89]<br><0.001 | 2.9<br>[2.90–2.93]<br><0.001 |
| **BC** |  | 0.6<br>[0.62–0.63]<br><0.001 | 0.32<br>[0.32–0.33]<br><0.001 | 1.1<br>[1.13–1.15]<br><0.001 | 0.7<br>[0.68–0.69]<br><0.001 |
| **CL** |  |  | 0.5<br>[0.51–0.52]<br><0.001 | 1.8<br>[1.80–1.83]<br><0.001 | 1.1<br>[1.08–1.10]<br><0.001 |
| **LC** |  |  |  | 3.5<br>[3.50–3.56]<br><0.001 | 2.1<br>[2.10–2.13]<br><0.001 |
| **CR** |  |  |  |  | 0.6<br>[0.59–0.60]<br><0.001 |

In males, post hoc pairwise comparisons indicated that all configuration pairs differ significantly from each other (*p* < 0.05). The GP configuration was rated least tolerable overall, whereas the PM configuration was rated most tolerable (Table 4).

**Table 4.**
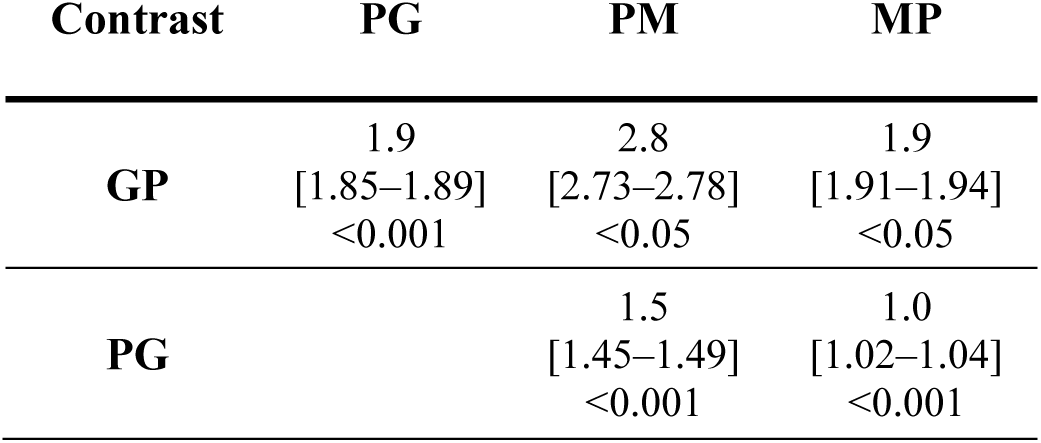
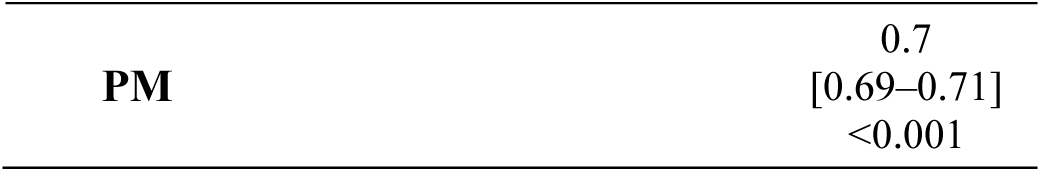
Pairwise CLMM contrasts comparing tolerability scores between pudendal configurations in male participants. Each cell shows the odds ratio (row vs. column), 95% confidence interval, and p-value. Odds ratios >1 indicate higher odds of reporting higher tolerability scores (less tolerable) for the row configuration relative to the column configuration.

**Table 5.**
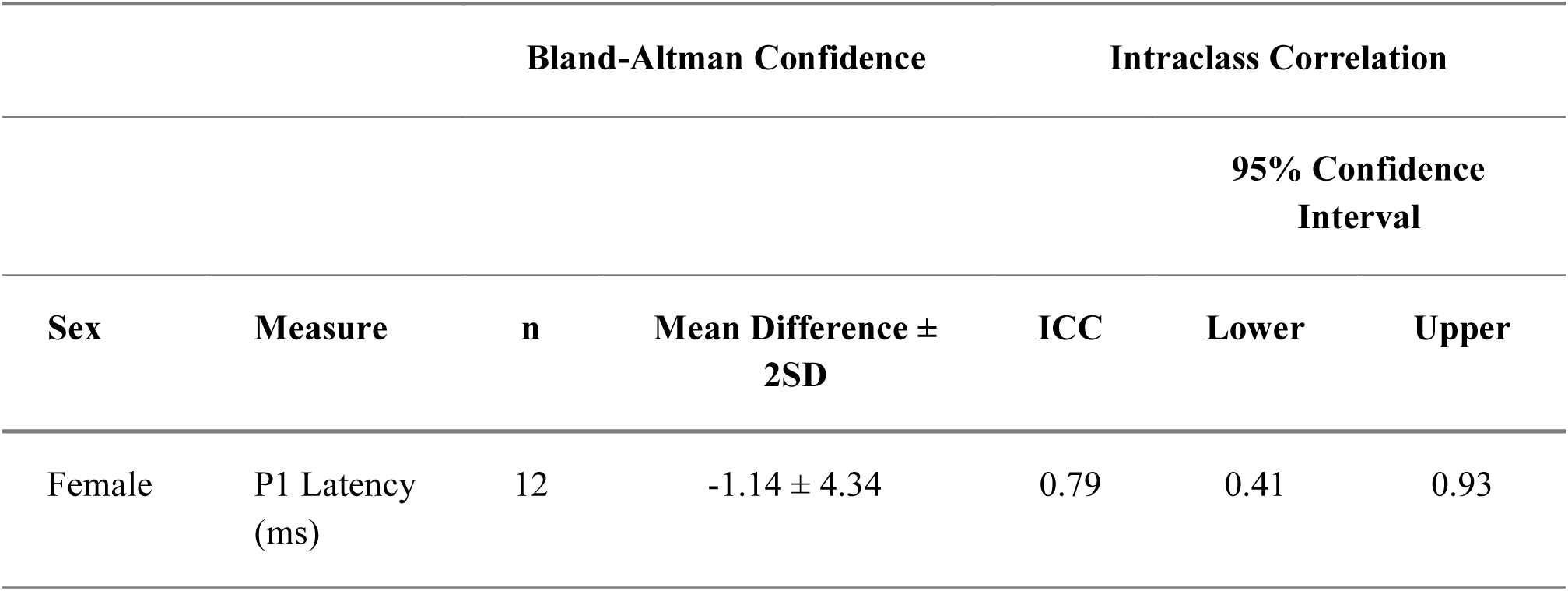
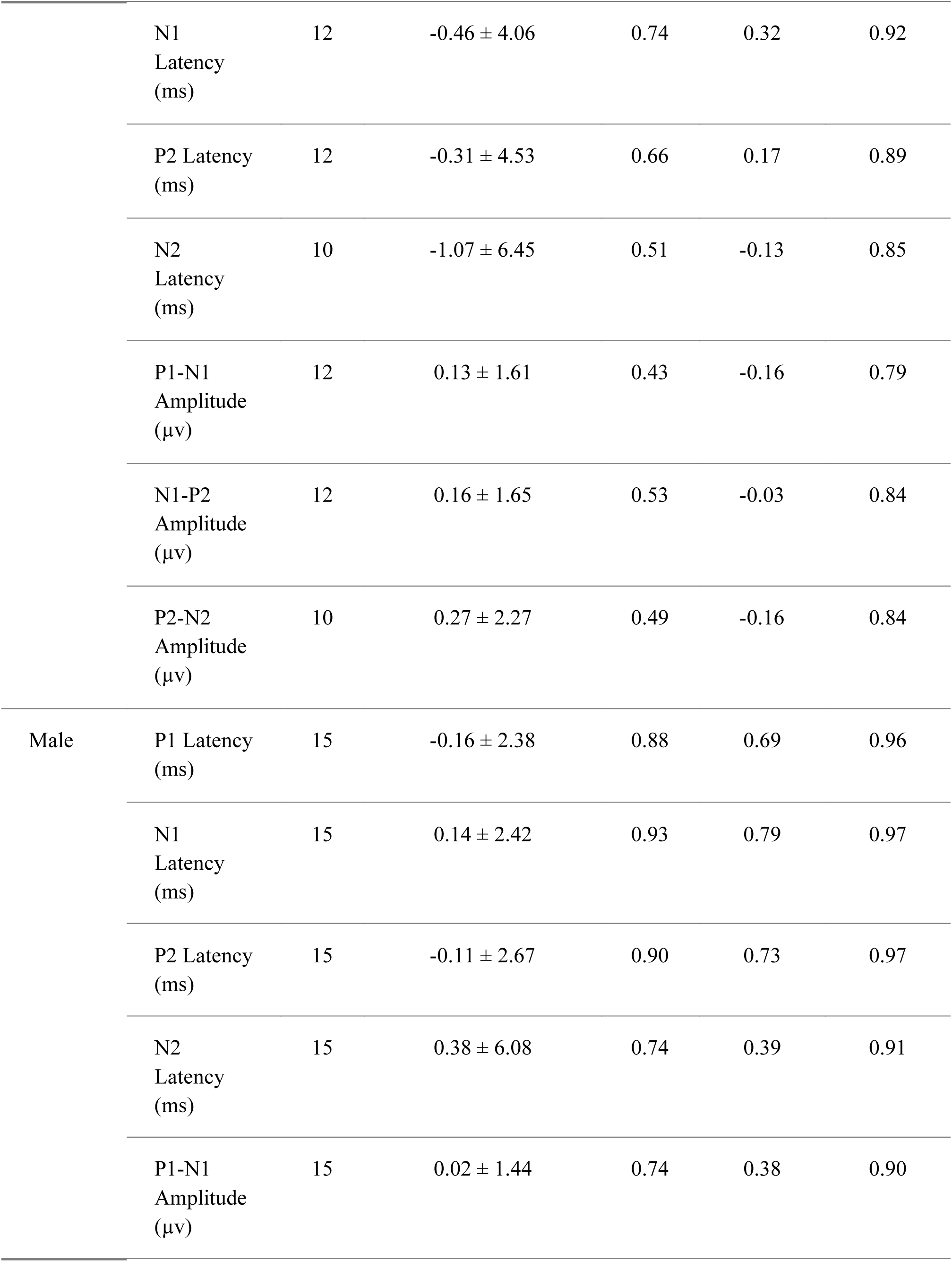
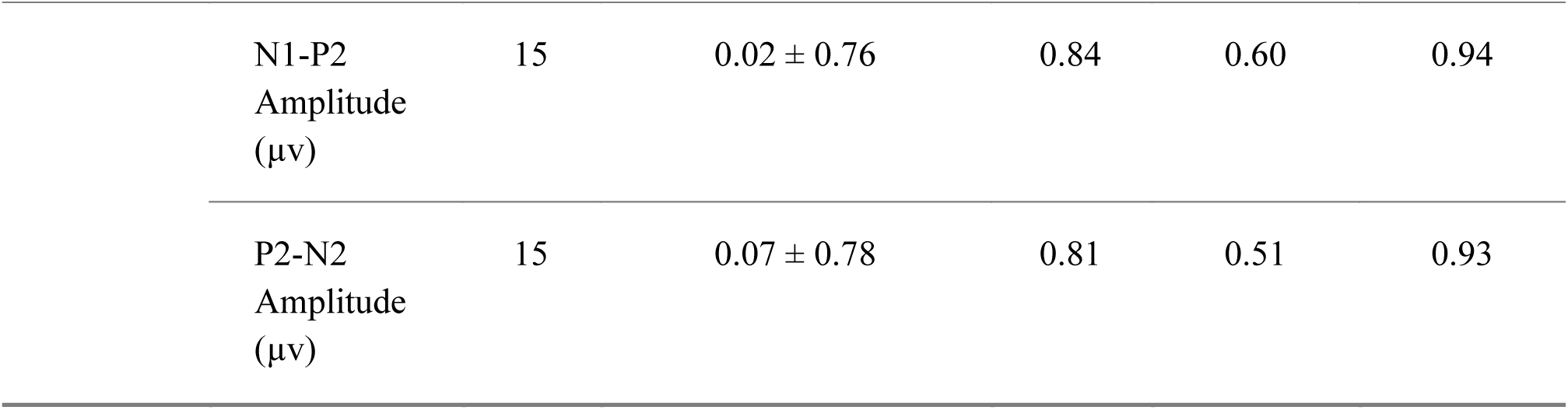
Test-retest reliability results for latency and amplitude measures.

Our combined CLMM revealed a significant interaction between nerve type and sex (odds ratio (OR) [95CI] = 11.0 [2.2, 54.8]; *p* = 0.004), indicating that the effect of nerve type on tolerability differed between females and males. Among females, tolerability scores were lower for tibial compared to pudendal stimulation, but the contrast was ten times less than that of the males (OR [95CI] = 0.03 [0.01, 0.2]; *p* < 0.001). Similarly, males also rated tibial stimulation as significantly more tolerable than pudendal stimulation (OR [95CI] = 0.3 [0.1, 1.2]; *p* = 0.047).

### 3.5 Response stability

Out of 227 trials across 42 participants, data from 167 trials could be fit to the exponential function with R^2^ ≥ 0.80. Five trials yielded τ_95_ values ≥ 2048 stimuli, so these were removed from this part of the analysis. From the remaining data, τ₉₅ values were calculated for each configuration, with each participant contributing one value per configuration. These values were then pooled across all participants and configurations. On average, we found that pudendal SEP responses stabilized at values well below 600 stimuli (females τ_95_ = 282, SD: 237, range: 17–1200; males τ_95_ = 324, SD: 206, range: 47–784).

For each of the pudendal SEP complexes, we compared the P2P_600_ and P2P_τ95_ amplitudes. The P2P_600_ amplitudes were consistently lower than the P2P_τ95_ amplitudes (Fig. 6), resulting in significant differences for the P1N1 complex (Δ = -0.3, t(16) = -4.5, *p* < 0.001, *d* = -0.3) and N1P2 complex (Δ = -0.2, t(17) = -3.0, *p* = 0.01, *d* = -0.2), while no difference was observed for the P2N2 complex (Δ = -0.1, t(17) = -0.7, *p* = 1.0, *d* = -0.05) in females. Similarly in males, the P2P_600_ amplitudes was consistently lower than P2P_τ95_ amplitudes for the P1N1 complex (Δ = -0.1, t(19) = -2.8, *p* = 0.02, *d* = -0.2), N1P2 complex (Δ = -0.1, t(20) = -2.1, *p* = 0.1, *d* = -0.1), and P2N2 complex (Δ = -0.1, t(19) = -2.6, *p* = 0.04, *d* = -0.2).

**Figure 6.**
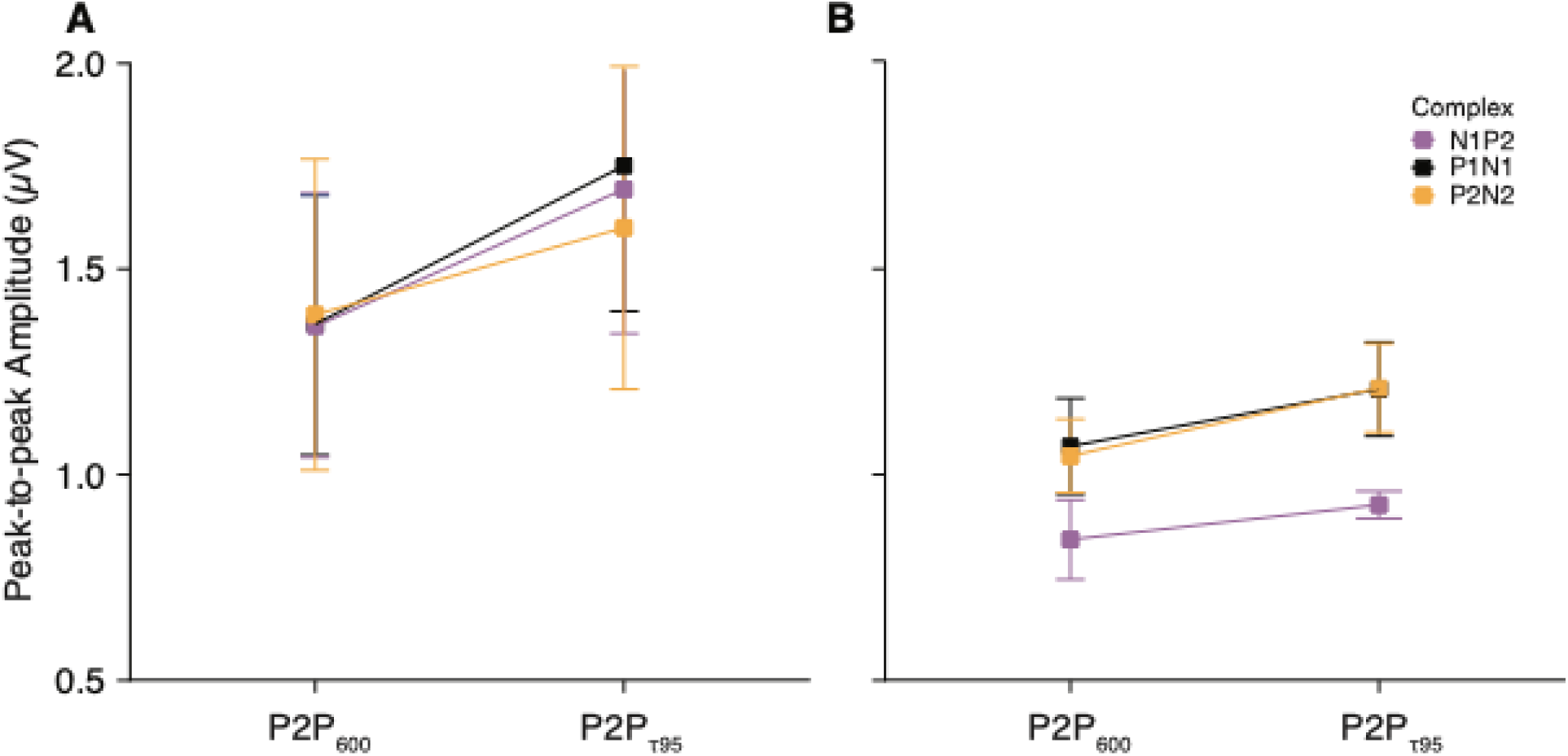
Comparison of the P2P_600_ amplitudes versus the P2P_τ95_ amplitudes across all three SEP complexes, shown separately for female (A) and male (B) participants. Across configurations, the P2P_600_ amplitudes were consistently lower than the P2P_τ95_ amplitudes.

Figure 7 presents the mean τ_95_ for the pudendal and tibial configurations across SEP complexes, separated by sex. There was no effect of electrode configuration on τ_95_ in either females (P1N1 complex: *p* ≥ 0.2; marginal R² = 0.1; f² = 0.1; N1P2 complex: *p* ≥ 0.2; marginal R² = 0.1; f² = 0.1; P2N2 complex: *p* ≥ 0.6; marginal R² = 0.03; f² = 0.03), or males (P1N1 complex: *p* ≥ 0.2; marginal R² = 0.1; f² = 0.1; N1P2 complex: *p* ≥ 0.4; marginal R² = 0.01; f² = 0.01; P2N2 complex: *p* ≥ 0.9; marginal R² = 0.001; f² = 0.001).

**Figure 7.**
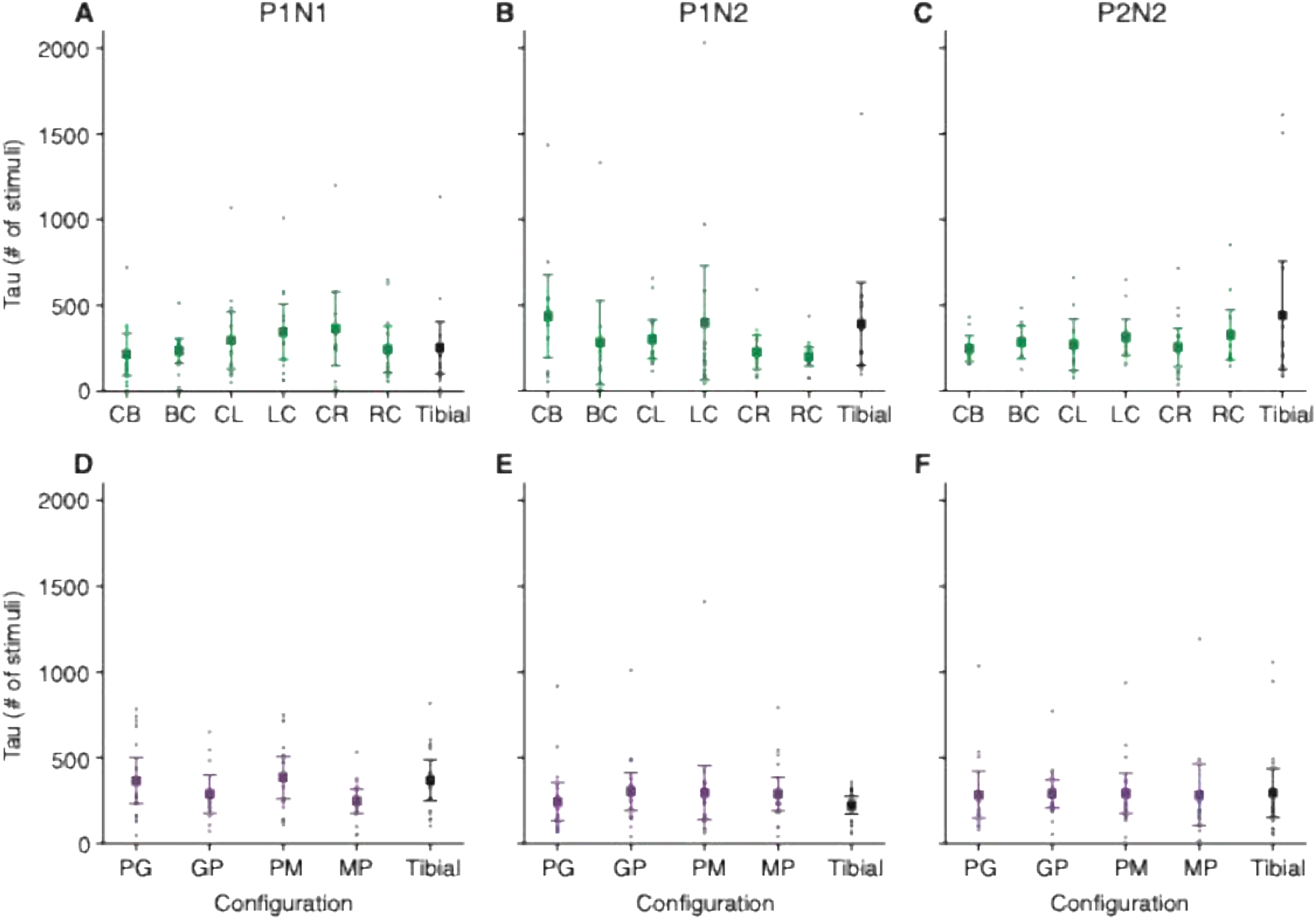
Mean τ₉₅ values with 95% confidence intervals for each pudendal electrode configuration across the three SEP complexes (P1N1, N1P2, P2N2), presented separately for female (A, B, and C) and male (D, E, and F) participants.

As a comparison, for the tibial SEP responses, the mean τ_95_ across all participants was 311 (SD: 249; range: 17–1132). There was no difference in τ_95_ between pudendal and tibial stimulation for any of the complexes (P1N1 complex: Δ = -41.7, t(28) = -0.8, *p* = 1.0, *d* = -0.2; N1P2 complex: Δ = -32.2, t(29) = -0.6, *p* = 1.0, *d* = -0.2; P2N2 complex: Δ = -74.8, t(29) = - 1.0, *p* = 1.0, *d* = -0.2).

### 3.6 Test-retest reliability

Twenty-seven participants (12 females, 15 males) were available for retesting. The mean interval between sessions was 84 days (SD: 34, range: 44–154). Three participants each had one trial excluded due to inability to tolerate stimulation at or above 2× PT.

In females, latency reliability ranged from moderate to good, with the lowest reproducibility observed for N2 (ICC [95CI] = 0.51 [−0.1 to 0.9], mean difference = −1.0 ms, SD: 3.2) and the highest for P1 (ICC [95CI] = 0.8 [0.4 to 0.9], mean difference = −1.1 ms, SD: 2.2). Males demonstrated generally higher reliability across latency measures (ICCs: 0.7–0.9; mean differences: −0.1 to 0.4 ms, SD: 1.2–3.0).

Peak-to-peak amplitude reproducibility was more variable, ranging from poor to moderate in females (ICCs: 0.4–0.5; mean differences: 0.1–0.3 µV, SD: 0.8–1.1) and moderate to good in males (ICCs: 0.7–0.8; mean differences = 0.02–0.1 µV, SD: 0.4–0.7). For all measures, mean differences between sessions did not differ from zero, indicating no systematic bias between visits. Bland–Altman plots are provided in the supplementary materials and show no detectable difference in agreement across test days for pudendal configuration or sex.

## 4.0 Discussion

The purpose of this study was to investigate the effects of stimulating electrode configuration and sex on pudendal SEP peak-to-peak amplitude, tolerability, response stability, and test-retest reliability in healthy adults. The specific electrode configuration used to stimulate the pudendal nerve did not affect peak-to-peak amplitude for any of the SEP complexes in either females or males. In contrast, tolerability scores varied across electrode configurations, with the CB configuration being rated as the least tolerable in females and the GP configuration being rated as the least tolerable in males. Participants generally rated pudendal stimulation as less tolerable than tibial stimulation, with females reporting a larger difference in tolerability of pudendal vs. tibial stimulation than males. On average, peak-to-peak amplitudes stabilized around 300 stimuli (females: 282, males: 324). Response stability did not differ across electrode configurations or between pudendal and tibial SEP waveforms. Finally, test-retest reliability was better for latencies than for peak-to-peak amplitudes, with narrower confidence intervals and higher ICCs overall. We found no evidence of systematic bias between sessions.

### 4.1 Physiological and practical implications of electrode configuration

The consistency of pudendal SEP amplitudes across configurations, despite differences in polarity and electrode placement, suggests that pudendal SEP waveforms are robust to electrode configuration variability. Electrodiagnostic guidelines often specify that the cathode should be placed proximal to the anode, in order to avoid anodal blocking, a mechanism by which a proximally placed anode may hyperpolarize nerve fibers and inhibit action potential propagation (Brindley & Craggs, 1980; Cruccu et al., 2008; Tavee, 2019; Toleikis et al., 2024; Vuckovic et al., 2008). Notably, most of the evidence supporting anodal blocking stems from animal models and motor or autonomic nerve stimulation (Brindley & Craggs, 1980; Tosato et al., 2007).

However, several investigations in humans, including intraoperative and clinical SEP studies, have found no evidence of anodal block, reporting comparable responses with cathodal and anodal stimulation (Allison et al., 2022; Dreyer et al., 1993; Kanbayashi et al., 2017; Kirshblum et al., 1998; Winkler & Stålberg, 1988). Our results similarly suggest that anodal blocking did not affect the pudendal peak-to-peak amplitude, as amplitudes were not significantly different between configurations where the anode was placed proximally or distally relative to the cathode in both males and females. However, other studies have shown that the effectiveness of anodal blocks is highly dependent on stimulus intensity, where higher intensities, smaller electrodes, and smaller inter-electrode distances are more likely to result in anodal blocking (Ahmed et al., 2020; Brindley & Craggs, 1980; Vuckovic et al., 2008). Nevertheless, our results suggest that the stimulation intensity and electrode configurations used to elicit pudendal SEPs were insufficient to elicit anodal block.

Moreover, electrode configuration affected tolerability, which is a critical consideration for the feasibility of pudendal SEP recording protocols, particularly given the sensitive tissues innervated by the nerve. Discomfort can increase participant burden and risk incomplete datasets due to skipped trials or early withdrawal.

In males, the GP configuration, where the anode is positioned over the glans and the cathode along the penile shaft, has been the most commonly used setup in previous pudendal SEP studies (Williams et al., 2024). This convention likely arose from standard electrodiagnostic recommendations that the cathode be placed proximally to avoid potential anodal block (Brindley & Craggs, 1980; Cruccu et al., 2008). However, as noted earlier, our findings and previous human studies suggest that anodal blocking does not influence pudendal SEPs under typical stimulation parameters. Given that both configurations produced comparable amplitudes, the choice between GP and PG can therefore be guided by participant comfort.

We found the GP configuration to be significantly less tolerable than the PG. The glans mucosa contains a dense network of Aδ and C-fiber free nerve endings, which makes it highly sensitive to electrical stimulation (Shih et al., 2013). Finite-element modeling has demonstrated that peak current density is concentrated directly beneath the electrode and decays with depth and distance, with higher local current density associated with greater discomfort in superficial tissues (Sha et al., 2008). Applying this principle, when current enters through the glans in GP, the resulting steep and focused electric field gradient likely induces stronger depolarization of superficial nociceptors, producing a sharper and more uncomfortable sensation. When the glans lies under the cathode (PG), stimulation still activates pudendal afferents, but shifting the anodal entry point to the penile shaft may produce a more distributed electric field before it reaches the glans. This could reduce the sharp, surface-concentrated activation of nociceptors observed when the glans itself serves as the anode, potentially explaining why participants rated PG as more tolerable. These results suggest that although GP remains the traditional configuration, its use likely stems from earlier recommendations intended to prevent anodal block. Given that our data showed no evidence of such effects, PG configuration could offer a more participant-friendly alternative while maintaining comparable SEP amplitudes and latencies to GP configuration. Future studies could consider adopting this configuration when feasible.

In females, tolerability patterns appeared more nuanced. Participants rated CB as the least tolerable configuration overall, yet the other bilateral configuration, BC, was rated as more tolerable than all unilateral configurations except CR. This suggests that bilateral stimulation does not inherently reduce comfort, despite engaging a broader sensory field through stimulating both branches of the pudendal nerve simultaneously. As with males in the GP configuration, we suspect that the worse tolerability scores for CB stem from strong electric field gradients near the clitoris, where the anode sits. This abrupt entry point likely leads to sharper sensory activation and a more uncomfortable experience, especially when compared to BC, where polarity is reversed. However, because electrode placement was participant-guided, we could not confirm the precise relationship between the anode and clitoral hood across participants. Small shifts in electrode position could have contributed to variability in tolerability scores, and future studies using standardized placement protocols are needed to clarify the relative contributions of polarity and electrode location to tolerability.

### 4.2 SEP responses stabilized well before the 600-stimulus mark

Our findings demonstrate that the cumulative average of the pudendal SEP waveform amplitudes reached a steady state, on average, at 282 stimuli in females and 324 in males, indicating that reliable responses can be obtained with far fewer trials than traditionally recommended. While conventional guidelines recommend averaging approximately 500 stimuli for cortical SEPs and up to 2000 for spinal recordings (Cruccu et al., 2008), our data, derived from τ_95_ modelling of response stability, support that stimulus count may be reduced without compromising signal stability in healthy adults. The ability to acquire robust SEP waveforms with fewer stimuli has practical value. In clinical and research settings, long-duration protocols may increase participant burden, which may in turn make it difficult for individuals to remain still or complete all required trials. However, it is important to interpret these results in the context of existing literature. Williams et al. (2024) reported that studies involving healthy participants and those with urogenital dysfunction typically used a median of approximately 250 stimuli, about half the number recommended by earlier guidelines. Our findings support the adequacy of these shorter protocols and demonstrate that reliable pudendal SEPs can be obtained with fewer repetitions in healthy participants. Future work should examine whether similar efficiencies can be achieved in clinical populations.

### 4.3 Latency measures show greater reliability than amplitudes

Williams et al. (2024), in a systematic review of 132 studies investigating pudendal SEPs across healthy and clinical populations, found that latency was reported in 115 studies (87%), whereas amplitude was reported in only 46 studies (35%). This emphasis on latency likely reflects its established diagnostic and methodological advantages. Latency abnormalities are sensitive indicators of conduction integrity and neurological recovery (Cui et al., 2015; 2019), and latency parameters show lower inter-subject variability than amplitude, making them more suitable for defining normal reference values (Miura et al., 2003). Consistent with this, our results demonstrated that latency measures also exhibited higher test–retest reliability than amplitudes across both sexes.

From a practical standpoint, these findings suggest that latency and amplitude serve distinct purposes depending on the experimental or clinical context. As latency reflects the timing of neural conduction, it provides a stable and interpretable metric for longitudinal or diagnostic assessments, particularly in clinical populations where monitoring changes in conduction delay or recovery over time is relevant. In contrast, amplitude is more variable between sessions but remains highly informative within a single recording period, where it can capture acute modulations in cortical or spinal excitability. Thus, amplitude measures are likely more valuable for same-day or cross-sectional studies investigating physiological mechanisms or short-term intervention effects, whereas latency offers a reliable outcome for repeated assessments over longer timescales.

### 4.4 Sex-related differences in pudendal SEPs

We found no sex-based amplitude differences. Previous studies have reported mixed findings, with some showing slightly larger early components in females, such as N15 (Mase et al., 2025) or N20 (Anazawa et al., 2023), whereas others reported greater P25-N30 amplitudes in females (Demura et al., 2024). Importantly, each of these studies also found no significant sex differences in several other SEP components. Earlier work by Ikuta and Furuta (1982) and Kakigi and Shibasaki (1991) similarly found higher SEP amplitudes in females across several components, including P27 and P45, though later magnetoencephalography studies observed no corresponding sex differences in source strength (Huttunen et al., 1999). This suggests that apparent amplitude differences may partly reflect biophysical factors, such as skull or cranial volume-conduction properties, rather than intrinsic neurophysiological variation. Taken together, these studies indicate that sex-related effects on amplitude are component-specific and inconsistent across studies.

In contrast, we found shorter pudendal SEP latencies in female participants (Table 2), consistent with previous studies reporting shorter somatosensory evoked potentials peak latencies in females than males (Huttunen et al., 1999; Ikuta & Furuta, 1998; Mase et al., 2025; Pelliccioni et al., 2014). Previous research supports the role of conduction path length in influencing SEP latency. For instance, Mervaala et al. (1988) demonstrated a strong correlation between height and median-nerve SEP peak latencies and recommended adjusting for height, age, and sex when interpreting normative data. However, Pelliccioni et al. (2014) included height as a covariate and still observed significantly shorter pudendal SEP latencies in females, suggesting that factors beyond conduction distance, such as anatomical or central conduction differences, may also contribute. Further studies that combine electrophysiological and anatomical assessments are needed to clarify the relative contributions of body size, peripheral conduction, and central processing to sex-related latency differences.

The observed sex differences in the reliability of SEP amplitudes and latencies are likely shaped by both anatomical and methodological factors. The pudendal nerve gives rise to multiple sensory branches, including dorsal clitoral or penile, posterior labial or scrotal, and perineal cutaneous nerves, with documented variability in branching patterns, fiber counts, and axon myelination along its course (Tunçkol et al., 2024). While such variability exists in both sexes, its impact may be more pronounced in females, given the smaller stimulation area and the potential for slight shifts in skin tension or electrode position within the vulvar region. This anatomical complexity increases the challenge of consistently activating the same fiber populations across sessions. Moreover, studies have shown that small changes in electrode placement near the clitoris (e.g., at 3- and 9-o’clock positions) can significantly alter SEP amplitude (Cavalcanti et al., 2007), supporting the idea that even subtle shifts in electrode position, angle, or pressure between sessions could contribute to increased variability. By contrast, male external genitalia provide a more standardized stimulation surface, which may facilitate more consistent fiber recruitment across sessions. To improve reproducibility, particularly in female participants, future studies should consider implementing strategies such as having the same trained investigator place the electrodes across sessions. This approach mirrors clinical practice, where a neurophysiologist is typically responsible for ensuring consistent electrode placement.

## 5.0 Conclusion

Collectively, the findings of this study provide a comprehensive evaluation of pudendal SEP amplitude, latency, tolerability, and test-retest reliability across different pudendal electrode configurations. Our findings demonstrate that pudendal SEP amplitudes remained consistent across different electrode configurations and highlight the importance of ensuring consistent electrode placement, particularly in female participants. These results may help inform the development of more feasible and reliable SEP protocols for future clinical and research applications.

## Data Availability

All data produced in the present study are available upon reasonable request to the authors.

## Acknowledgements

We sincerely thank the participants for their time and commitment to take part in this study. We are also grateful to Megan Henry, Lauren A. Platz, Julian Lui, and Oscar Ortiz for their support in data collection and analysis.

## Declaration of Generative AI and AI-assisted technologies in the writing process

During the preparation of this work, the author occasionally used *ChatGPT (OpenAI, GPT-5)* to improve phrasing and clarity. All substantive content, analysis, and interpretation were written by the author. The final text was thoroughly reviewed and edited by the author, who takes full responsibility for the content of the publication.

