## Supplementary figures and images for "Pudendal somatosensory evoked potentials – A standardized assessment for males and females"

### Supplemental File

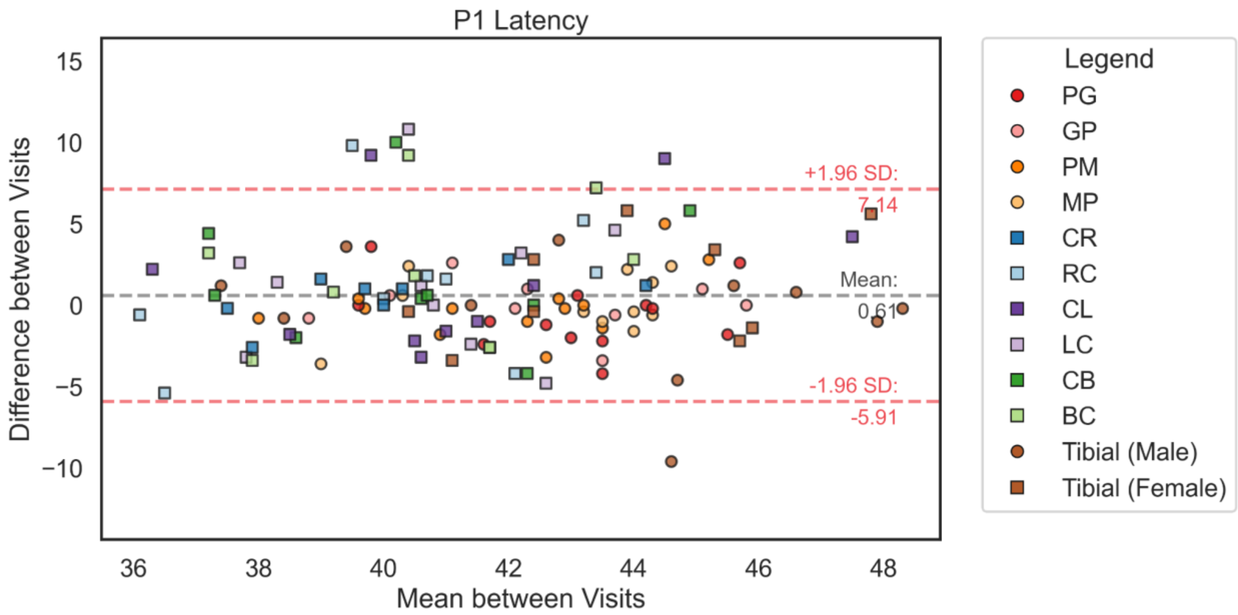

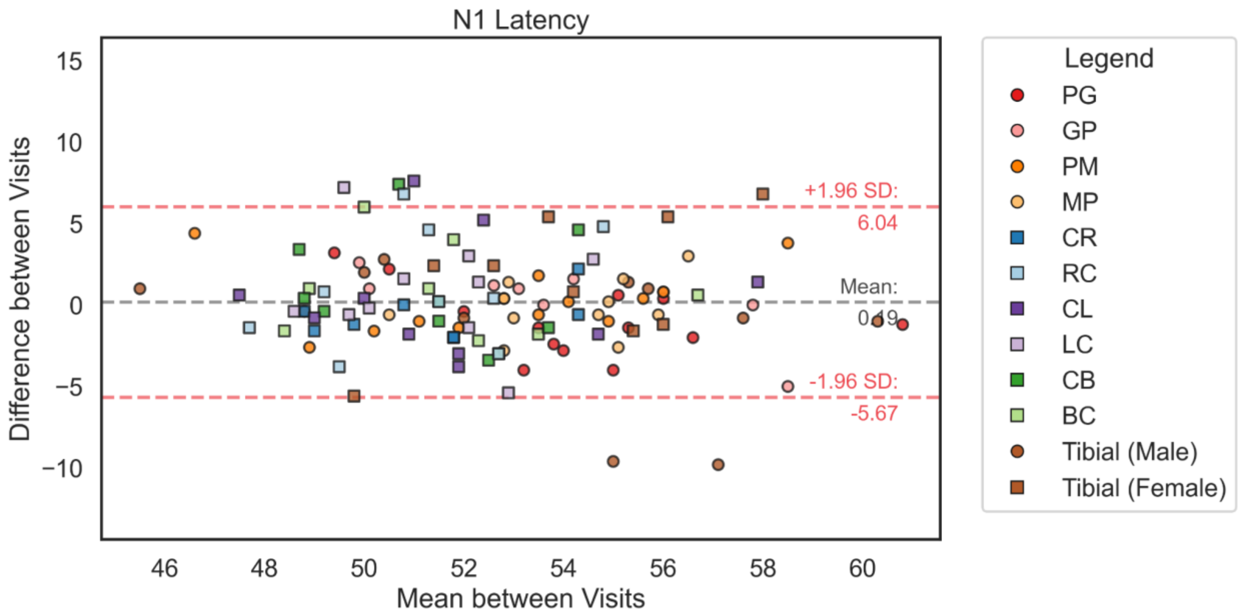

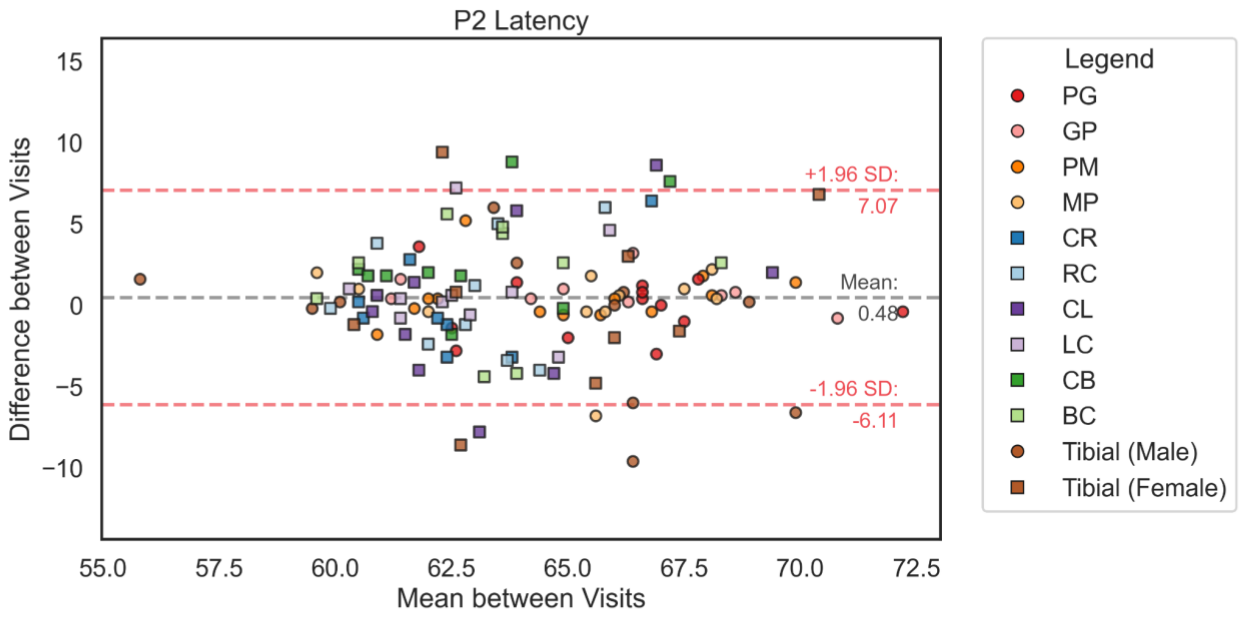

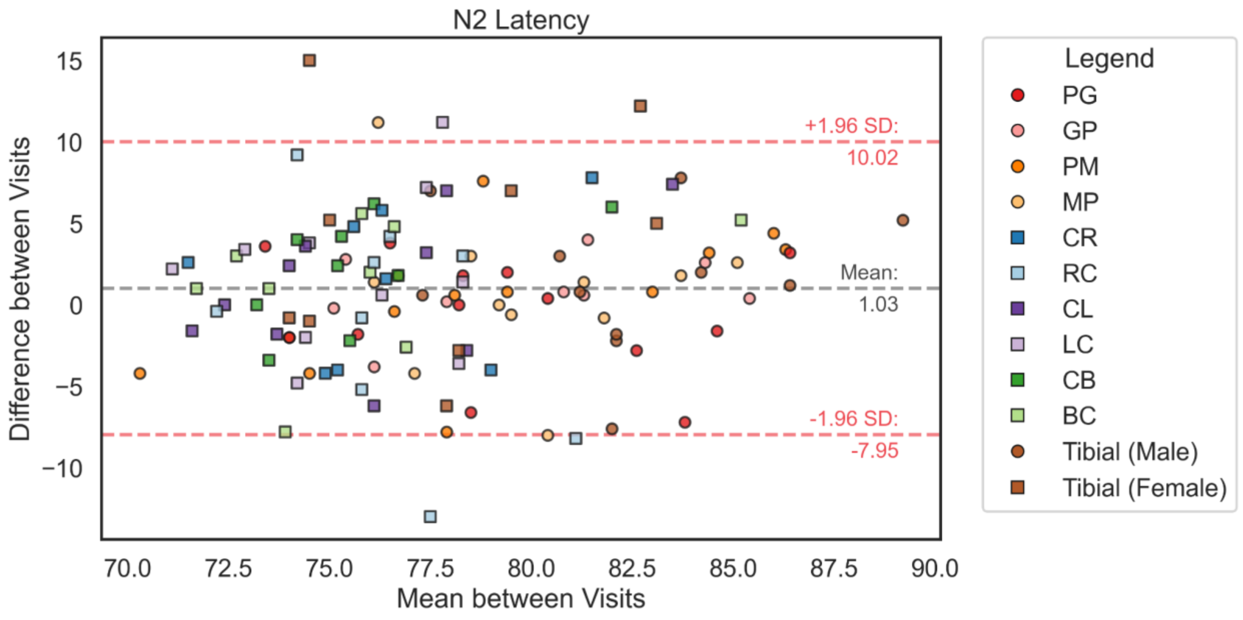

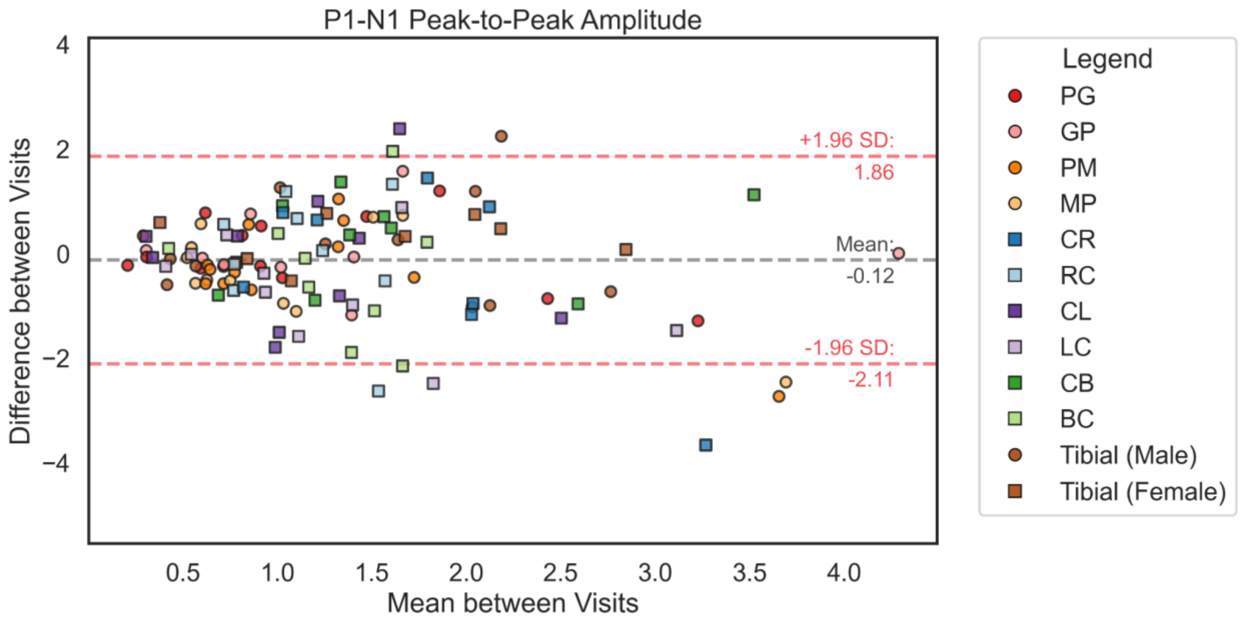

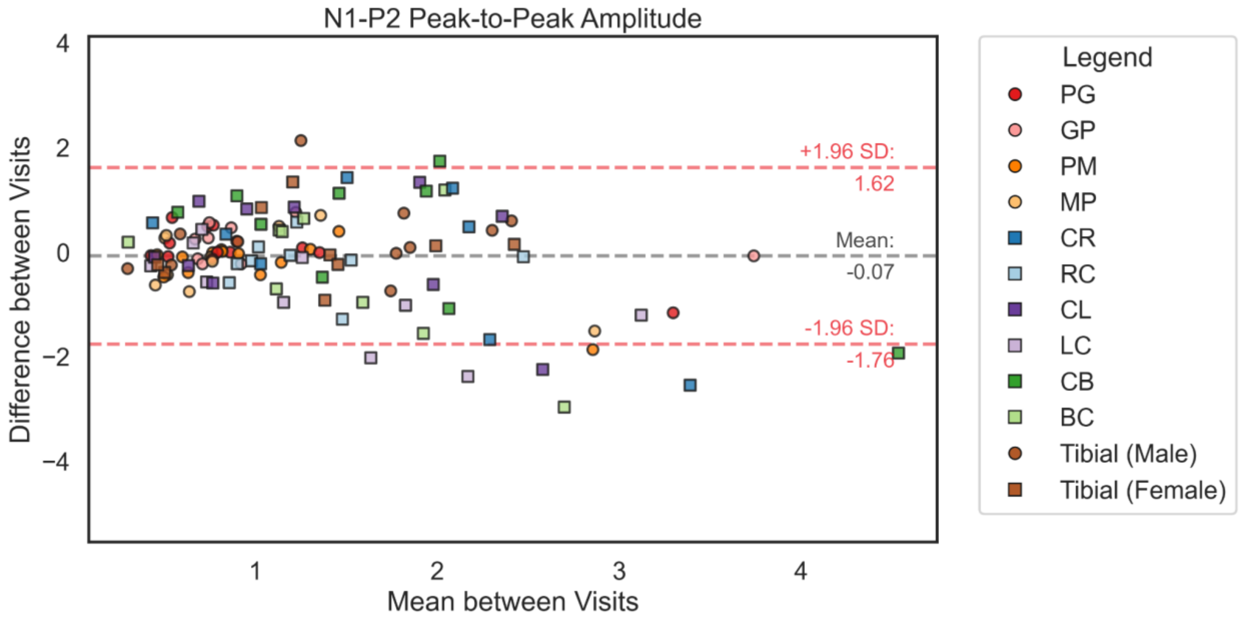

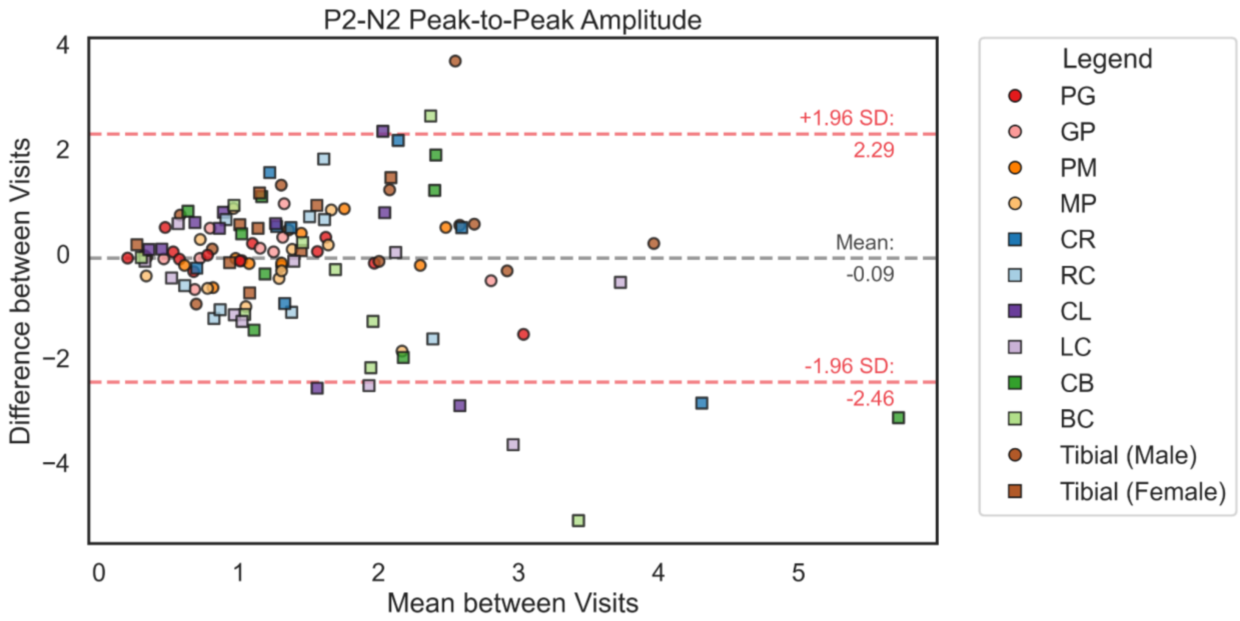
